# Barriers to access to HIV care services in host countries: views and experiences of Indonesian male ex-migrant workers living with HIV

**DOI:** 10.1101/2022.04.24.22274022

**Authors:** Nelsensius Klau Fauk, Alfonsa Liquory Seran, Christopher Raymond, Roheena Tahir, Paul Russell Ward

## Abstract

This study aimed to understand barriers to accessing HIV care services in host countries among Indonesian, male, former (returned) migrant workers living with HIV. The study utilised a qualitative design employing in-depth interviews to collect data from twenty-two returned migrant workers from Eastern Indonesia, recruited using the snowball sampling technique. A qualitative data analysis framework was used to guide a step-by-step analysis of the findings. Findings demonstrated that limited host-country language proficiency, lack of knowledge regarding healthcare systems in host countries and having ‘undocumented’ worker status were barriers to accessing HIV care services. Data also revealed unavailability of HIV care services nearby migrants’ work locations, long distance travel to healthcare facilities, and challenges in accessing public transportation as barriers that impeded their access to the services. Other factors limiting the participants’ access to HIV services were identified as the transient and mobile nature of migrant work requiring frequent relocation and disrupting work life stability. Additionally, in lieu of formal HIV services, many participants self-medicated by using over-the-counter herbal or ‘traditional’ medicines, often because of peer or social group influence regarding selection of informal treatment options. Recommendations arising from this study demonstrate the need to improve pre-departure information for migrant workers regarding healthcare system and access procedure in potential host countries. Data from this study also indicate that social services should be available to assist potential migrants to access legal channels for migrant work overseas, to ensure that Indonesian migrants can safely access healthcare services in the countries for which they are providing migrant labour. Future studies to understand barriers to accessing HIV care services among various migrant groups living with HIV are warranted to build evidence for potential social policy change.

## 1. Introduction

Migrants, who are mostly from low/middle income and often HIV-endemic countries, are increasing population groups in many developed countries (1, 2). In terms of migrant workers in particular, the International Labour Organisation has reported an estimated 169 million international migrant workers globally in 2020 (3). Indonesia is one of the countries that supply migrant workers to other countries, with an estimated nine million migrant workers from Indonesia working in more than twenty-five countries throughout the world (4, 5).

Migrant workers are defined as people who migrate from one country to another for work purposes and without an intention to stay permanently in the country where they work (6, 7). Migrant workers are a group at high risk for HIV transmission and its consequences (8, 9), with studies showing higher HIV prevalence and higher frequency of delayed HIV diagnosis among migrants than the general populations (10-13). Migrant workers are also reported to experience delayed entry or linkage into HIV services and have poorer HIV-related health outcomes compared to the general population in host countries (14-16).

Numerous studies have reported that migrants living with HIV (MLHIV) face numerous barriers that hamper their access to HIV care services. These can be conceptualised as individual, interpersonal, community/social and policy related barriers. Individual-level barriers to HIV services include fear of deportation and negative social consequences of HIV status disclosure, such as stigma, discrimination, social isolation and job loss (17-20). The fear of deportation is reported mainly among migrants who are undocumented, since their access to the services may lead to the discovery of their undocumented status by authorities (9, 21-24). Fear of negative side effects of antiretroviral therapy (ART) is also another factor that impedes access to HIV services for MLHIV (20, 25, 26). Language difficulties and a lack of knowledge and education about HIV, HIV care services and unfamiliarity with healthcare systems in host countries are also individual-level barriers to the access of MLHIV to HIV care services or ART (18, 23, 26-29). Interpersonal barriers to services include a lack of social support from families or friends, who are aware of their HIV status, to link them to HIV care services (30). Stigma and discrimination from others within communities and from healthcare providers in healthcare facilities are community/social barriers that impede access for MLHIV to HIV services (18, 20, 31). Some studies have also reported policy-level barriers such as immigration policies in host countries which do not cover migrants for free access to HIV care services and exempt them from national health insurance (26, 30, 32, 33).

Despite a range of different level of barriers to access to HIV care services by migrant populations in general who are living with HIV as reported in the aforementioned studies, most of these studies were conducted in developed countries, such as in Europe, USA, Canada and Australia (8, 9). There is a paucity of evidence about barriers to access of HIV services for MLHIV in developing countries. This paper aims to fill in this gap in knowledge by exploring the views and experiences of Indonesian male ex-migrant workers who are living with HIV about barriers to their access to HIV care services in host countries where they worked. Understanding barriers to access to HIV services by MLHIV are critically important to inform policy and intervention programs in order to develop appropriate access to HIV services for them in the future.

## 2. Methods

To support the explicit and comprehensive reporting of this qualitative study, consolidated criteria for reporting qualitative studies (COREQ) checklist was used to guide the report of the methods section (S1 Fig) (34).

### 2.1 Study setting

This study was carried out in Malaka and Belu districts, East Nusa Tenggara province, Indonesia. Malaka district was part of Belu district before the separation in 2012. It shares the border with East Timor in the East, *Timor Tengah Utara* and *Timor Tengah Selatan* districts in the West, and Belu district in the North (35, 36). Malaka covers an area of 1,160,63 km^2^ and has a total population of 171,079 people comprising 83,492 male and 87,587 female (35, 36). It has 12 sub-districts 1 hospital, 17 community health centres, 25 sub-community health centres, 5 private clinics, and 82 village health posts or village maternity posts (35, 36). Belu district shares a border with East Timor in the East, *Timor Tengah Utara* district in the West and Malaka district in the South (37). Belu has a total area of 1,284,94 km^2^ and a total population of 204,541 people including 100,922 male and 103,619 female (36, 37). Belu consists of 12 sub-districts and has 2 private and 1 public hospitals, 17 public health centres, 21 sub-public health centres, 48 village maternity posts, 23 village health posts and 5 private clinics (36, 38). In terms of migrant workers, it is estimated that there are 4200 Indonesian migrant workers from East Nusa Tenggara province (including Malaka and Belu) who are working overseas (39). To the best of our knowledge, there have never been any studies exploring barriers for Indonesian migrant workers, including the ones living with HIV, to accessing health care services in general or HIV care services in particular in host countries. Because of feasibility, familiarity and the potential of undertaking the current study successfully, Malaka and Belu districts were selected as the study settings.

### 2.2 Participant recruitment and data collection

This study utilised a qualitative methodology employing in-depth, in-person interviews to collect data from Indonesian male ex-migrant workers living with HIV, having returned from working abroad. Participants (n=22: 13 from Malaka and 9 from Belu) were recruited using the snowball sampling technique. Prior to recruiting participants, directors of several government public health centres and a public HIV clinic were approached by a member of the research team (ALS, a female public health researcher by training) to request their assistance in distributing study information sheets to potential participants who had accessed health services in their facilities. Following a formal agreement with these facilities, study information sheets were posted on information boards and distributed to potential participants by the receptionists in these healthcare facilities. Some potential participants texted the field researcher and asked for a meeting to clarify questions prior to their decision regarding participation. Potential participants who contacted and confirmed their willingness to be part of the study were then recruited for an interview. These initial participants were subsequently requested to disseminate the study information sheets to their eligible friends and colleagues. Study participants were selected based on the following inclusion criteria established by the research team: (i) aged at least 19 years old or greater, (ii) diagnosed as HIV positive, and (iii) having an HIV status diagnosed either in Indonesia prior to disembarking abroad for work or having been diagnosed subsequently while working abroad in the host countries. Three potential participants were excluded as their HIV was diagnosed subsequent to their return to Indonesia after working abroad, and thus had no experience of HIV care seeking in the host country. Once the participant roster was finalized, none of the confirmed participants withdrew from the study. None of the participants had established relationships with the researchers prior to inclusion in the study. A participant roster of 22 male ex-migrant workers from Malaka and Belu districts living with HIV participated in this study.

The participants’ age ranged from 25 to 48 years, duration of work abroad as migrants ranged between one and four years. The countries where they migrated to for work were Malaysia, Taiwan, Thailand and Hong Kong, China. The majority of the participants graduated from junior high school, followed by senior high school and elementary school, while a couple of persons were elementary school drop outs. Most of the participants had been diagnosed with HIV within a period of 6-10 years and a few were diagnosed with HIV for 1-5 years. Most participants (n=18) had been diagnosed with HIV prior to migration to work overseas, while a few were diagnosed with HIV in the host countries. All the participants were on ART when the study was conducted.

Data collection was carried out from December 2020 to February 2021. One-on-one and face-to-face in-depth interviews were carried out in a private room at the healthcare facilities on the day the participants accessed antiretroviral medicines or referral letter at the healthcare facilities. Both interviewer and participants abode to COVID-19 guidelines by maintaining a physical distance of 1.5 metres and wearing facemasks during the interviews. The time and place for the interviews were mutually agreed upon by both researcher and each participant. Prior to the interviews, the participants were again informed about the purpose of the study and the voluntary nature of their participation. They were also informed about their rights to withdraw their participation at any time for any reasons without consequences. They were assured that information or data they provided in the interviews would be made anonymous by assigning a pseudonym for each transcript. Interviews were performed in Bahasa Indonesia and only the researcher and each participant were present in the interview room. Interviews were recorded using a digital audio recorder. The researcher also took written notes during the interview when deemed necessary. The interviews ranged from 40 to 60 minutes in duration. Interviews were focused on several key areas, such as the migration process or how the participants got into the host countries, what kind of job they engaged in, their experiences with the language in the host countries and their knowledge about and experiences with health care services in the host countries. The interviews also explored the participants’ views about the availability of HIV care services in the areas where they stayed and worked, their experiences with public transportation to healthcare facilities, how they dealt with their health condition or HIV infection in the host countries, including the use of treatment other than ART, and what kind support they received from other people or friends or fellow migrant workers. Interviews and recruitment of the participants ceased when the researchers felt that data or information provided by the participants had been rich enough to answer the research questions and objective. The richness of the data was decided based on data saturation which was indicated in the similarity of answers or responses of the last few participants to those of previous participants (40). No repeated interviews were conducted with any participants. Participants were offered an opportunity to read and correct their interview transcript after the transcription but none required to do so. The participants were assured that information they provided would remain confidential and anonymous, and a pseudonym was assigned to each interview transcript. All participants signed and returned informed consent on the interview day prior to commencing the interview.

### 2.3 Data analysis

All the audio recordings were transcribed verbatim into coding sheets before the complete data analysis was performed. The comprehensive data analysis was carried out manually by the first author (NKF) and in Bahasa Indonesia to retain the cultural and social meanings of information provided by the participants (41). For the purpose of publication, a research team member (NKF), who is fluent in both Bahasa Indonesia and English, translated the quotes into English. The translation was then checked by other authors to maintain its accuracy and clarity. A qualitative data analysis framework (42, 43) was used to guide this data analysis process. This framework suggests five steps of qualitative data analysis: (i) familiarisation with the transcripts through reading them repeatedly, breaking down the information in each transcript into chunks of information and providing comments and giving labels to them; (ii) identification of a thematic framework through listing recurrent ideas, concepts and issues identified from the transcripts, which were then used to form the thematic framework; (iii) indexing the transcripts through creating open codes to each individual transcript, followed by creating a close coding through which similar or redundant codes were identified and collated to reduce the numbers of codes. Codes that formed a theme were grouped together under the same theme; (iv) charting the transcripts through reorganising the themes and their codes, that had previously been created, in a summary chart, and then comparisons of the data within and across interviews were performed; and (v) finally, mapping and interpretation of the data the entire data (43, 44).

Ethics approval for this study was obtained from the Health Research Ethics Committee, Duta Wacana Christian University, Indonesia (No. 1005/C.16/FK/2019).

## 3. Results

### 3.1 Language barriers, lack of knowledge of healthcare systems in host countries and being undocumented migrant workers

A lack of language proficiency was reported as influencing the participants’ intention and ability to access HIV care services in host countries. The participants expressed concerns that language barriers impeded their ability to explain their personal health issues, which also contributed to a lack of self-confidence when speaking with healthcare professionals in host countries. Thomas who spent a couple of years as a migrant worker abroad and remarked: ‘…*initially I intended to access ARV medicines, but the problem was that I did not know the language. I did not know how to tell the doctor or nurse*.’ Similarly, a lack of knowledge about healthcare systems in host countries hindered their access as they were unaware of any specific procedures to attain HIV care services:

> *“I had made up my mind before I left Indonesia and intended to access the [ARV] medicines, but when I was there [name of the country where he worked] I was not brave enough to look for and meet doctors because I didn’t know their language. I didn’t know how to explain what I was going through. Another concern was that I didn’t even know where to start. For example, here [in Indonesia] the procedure is that you just need to go to a public health centre and the nurse will examine your condition and refer you to the HIV clinic. But I didn’t know the procedure over there, so I didn’t make any efforts to access the [ARV] medicines while I was there” (Ali)*.

The condition of not having complete, legal documentation or being an undocumented migrant worker was another influencing factor for some participants (n=5) to access HIV care services in the host countries. These participants explained that they tried as much as they could to hide from authorities to avoid getting caught, which would have potentially led them to being detained or deported back to Indonesia. Such conditions were acknowledged as causative factors in their unwillingness to seek out HIV care or other health service, out of fear of reprisal. The following narrative of a participant who worked overseas for a few years reflects on his experience:

> *“At that time, I brought some medicines [ARV] from here [Indonesia], and after taking them all I didn’t try to access the medicines over there because I was afraid of getting caught by the authorities. I didn’t have documents; I was an illegal migrant worker. I was afraid that if I looked for the medicines, then doctors or nurses may ask for some documents and that would be a problem for me. I could get caught and jailed or deported. Sometimes, authorities such as the police went to check for illegal people [migrant workers] in the plantation areas. I remember some friends of mine and I had to run and hide in the forest a few times because of that”* (Isto).

### 3.2 Unavailability of HIV care services close to working sites

The unavailability of HIV care services or antiretroviral therapy in the sites or plantation areas where they worked was a barrier for migrant workers living with HIV to access these services. Some participants who were already on antiretroviral therapy prior to migrating abroad commented that they initially had the intention to access HIV care services in the host countries they moved into in order to adhere to the treatment: ‘*I wanted to access the [ARV] medicines over there but the medicines were not available*’ (Yos). However, they did not access them due to the unavailability of reliable HIV services at the workplace as reflected in the following story:

> *“In the plantation area where I worked, HIV care services or treatment were not available. After two months I arrived there [the place where he worked in Malaysia] I tried to find information about antiretroviral therapy. I was told that it was not available in the district. So, after I finished the [ARV] medicines I brought from here [Indonesia] I didn’t access the medicines because they were not available in the place where I worked. The nurse here gave me the medicines for two months and I planned to access the medicines when I arrived in XXX [name of the country where he worked], but I didn’t access because the medicines were not available. So, after two years working there, I started to feel that my physical condition was getting weaker” (Anton)*.

Long-distance travel requirements to general healthcare facilities or HIV clinics were other identified barriers that impeded participants’ access to HIV care services. Participants who shared their stories about the unavailability of nearby services in districts where they worked further explained that they received information about the availability of services in another district: ‘*I looked for information about HIV care services. I had a local friend who helped me find out about the services which were available in another district*’ (Melki). However, none of them accessed the services due to the long-distance travel, as mentioned by Gabriel, ‘*I knew about a hospital that provided the [ARV] medicines but it was too far away in another district, so I didn’t access it*’. In addition to the long-distance travel, a lack of public transportation and the fact that the participants did not have friends or relatives living in cities where the services were available, to help them access the services or with whom they could stay, were also underlying reasons that demotivated them to access services:

> “*When I first arrived in XXX [name of the country where he worked], I checked on the internet about HIV clinics so I knew. However, I worked in an oil palm plantation which was far from the city. The distance from the plantation to the city where there was an HIV clinic was very far. The problem was that there was no public transportation in the plantation area, so it was very difficult to get to the city. Besides, I didn’t know anyone or friends or relatives in the city, so at that time I thought if I went to the city then who would help me to get the access to HIV clinic and the [ARV] medicines, and where I should sleep because I believed I couldn’t go back to the plantation area on the same day, there was no transportation. Therefore, for almost three years I worked there I didn’t access the medicines”* (Kobus).

### 3.3 Mobile, transient nature of the work

The transient nature of their work, requiring them to move from one plantation area to another, was another underlying barrier that hindered participants from accessing HIV services. A few participants described how mobility in their work influenced them to dismiss considering seeking HIV care due to challenges arising from a constant change in environment and locales, making it difficult for them to adjust and adapt and find appropriate HIV support in new places. This is reflected in the following quotes from two migrant workers spending a few years circulating through various construction sites and plantation areas:

> *“I worked in XXX [name of the country where he worked] for two years as a construction worker. I found out that I had HIV in the second year. Initially, I had a problem with my throat; it was painful, and I couldn’t swallow food so I went to get a check-up with the doctor. After he checked my mouth, he asked me whether I would agree for him to also test for a virus called HIV and I said ‘yes’. After the test he told me that I have the virus in my blood. Then he gave me medicines for one month and asked me to go back to meet him afterwards but I didn’t go back because I had already moved to another place for construction work which was far away from the doctor’s office. We [construction workers] regularly moved from one place to another every two to three months. So, at that time I didn’t really think about accessing the services*” (Nelis).
>
> *“I worked on an oil palm plantation for three years in XXX [name of the country where he worked]. We regularly moved from one plantation area to another, which was often across multiple districts. It depended on the work and on our boss. If the boss said we had to move to another plantation area for a few months, then we would go. I knew about my HIV status before I went there, and I was on antiretroviral therapy medication before I arrived. I was thinking of continuing the treatment over there but it seemed impossible because my working situation was not supportive for me to access the medication as I regularly moved from one place to another. So, I didn’t even try to look for information about the services. I felt sick during the third year and that is why I decided to come back here [Indonesia] and restarted the treatment”* (Sius).

### 3.4 Self-treatment and the use of herbal and traditional medicines

The use of over-the-counter packaged herbal medicines was indicated as an influencing factor for participants’ access to HIV care services. Several participants (n=7) described that packaged herbal medicines were available over-the-counter at supermarkets or could be ordered online, which they then self-medicated in lieu of standardised ARVs to treat their HIV while working overseas. One of them stated “…*there are many supermarkets selling herbal medicines, so I bought and used them for the treatment of my HIV while I was in XXX [name of the country where he worked]*” (Goris). Participants also commented that herbal medicines were easier to acquire compared to standard ART, and could be purchased without the need for prescriptions from a medical doctor:

> *“I consumed packaged herbal medicines which I bought from supermarkets. Sometimes I ordered them online, but those are a little bit expensive. They were easier to access and I didn’t need any prescription from a medical doctor to get them. It is not like ARV medicines where everybody has to go through a lot of procedures and tests to be able to access them. So, I didn’t think of accessing ARV medicines. I worked nearby a hospital for more than one year and I knew about HIV care services there, but I didn’t access them. I took herbal medicines for nearly two years and then switched to [ARV] medicines once I came back here [to Indonesia]. I continued the treatment using herbal medicines for six months here [in Indonesia] because I bought a few bottles [while abroad] and brought them with me [to Indonesia]. Once I finished them all, I restarted the ARV medicines*” (Primus).

Similarly, the use of traditional medicines for HIV self-treatment influenced participants’ access to HIV care services in the host countries where they worked. Several commented that they brought traditional medicines from home and continued using them for HIV treatment while working overseas: *‘I started taking the [traditional] medicines here and brought them with me so I continued taking them over there [the country where he worked]*’ (Joni). Some respondents described that they made traditional medicines by themselves while working overseas, made of roots, leaves and the bark of plants which they could easily find in plantation areas. Meanwhile, others stated that they used traditional medicines obtained from local traditional healers nearby the places where they worked or lived:

> *“I brought a plastic bag of traditional medicines from here [Indonesia] given to me by a traditional healer in my village. But when I had finished taking them, I then made the medicines myself. I used roots, leaves and the bark of plants to make them. I found those herbal ingredients around the plantation area where I worked [abroad]. I harvested them, cleaned them up and boiled them, and then drank the infusion and also bathed myself with the infused water. A traditional healer here told me the names of the plants I should use, and how to mix them up so I knew*” (Nestor).
>
> “*One day health workers came to the plantation area and sampled our blood to check for malaria. After a week I was told that I didn’t have malaria, but that I had HIV. I knew some local people, so I asked them about traditional healers and they told me about a traditional healer nearby; he was the one who gave me the traditional medicines. I felt fine, so I didn’t think of taking ARV medicines. I was told by the health workers about [ARV] medicines from doctors but I didn’t access them. I continued taking the traditional medicines until I came back here [to Indonesia]” (Marsel)*.

### 3.5 Social influences

Social influence from friends played a significant role in influencing the participants’ access to HIV care services in the host countries. Some participants commented that their friends or other Indonesian migrant workers encouraged the use of packaged herbal medicines or traditional medicines. Okto, a young man in his mid-twenties, mentioned: *‘at that time another friend of mine and I tested positive for HIV. After the diagnosis, our friends encouraged us to use traditional medicines or packaged herbal medicines because they said these medicines are more effective for HIV treatment*.*’* The participants also acknowledged that their fellow migrant workers helped them to access traditional medicines from local traditional healers located in nearby communities:

> *“After I was diagnosed with HIV over there [the country where he worked], I asked a friend of mine to help me access HIV treatment, and he told me that he knew a traditional healer who could give [traditional] medicines. So, we both went to that traditional healer, and I got the medicines from him. I took the medicines from that traditional healer for about a year and didn’t access the medicines [ART] from doctors” (Sebas)*.

Peer influence was responsible for propagating misleading information about the potential negative consequences of disclosing HIV status as a foreign migrant worker to healthcare professionals in the host countries. Some participants described how their fellow migrant workers asked them to hide their HIV status from healthcare professionals as it would likely be reported to authorities, with a potential consequence that local authorities would investigate and detain them. The narrative of a participant who worked overseas for a few years reflected how peer influence through misleading or misinformation discouraged him from accessing HIV care services and encouraged the use of non-standardised, traditional, or packaged herbal medicines:

> *“I worked in XXX [name of the country where he worked] for a few years. I knew my HIV status before I went there, and I was already on medication [ART] here [in Indonesia]. I stopped the medication once I left for XXX [the country where he worked]. After two and a half years working there, I felt that my physical strength had reduced, and I was easily getting sick. So, I talked to a [Indonesian] friend of mine about my intention to access [ARV] medicines but he said that if the doctors were to know about my HIV status, they would report it to the police and I could be detained in prison. So, the solution was to purchase herbal medicines from supermarkets. A friend of mine bought those medicines for me at the beginning …” (Agus)*.

## 4. Discussion

Migrant populations are one of the vulnerable groups to HIV transmission. Previous studies in different settings have shown that HIV prevalence among migrants is higher than in general populations (10-13, 45, 46). This study explored the views and experiences of Indonesian male ex-migrant workers regarding barriers to accessing HIV care services in host countries. A lack of language proficiency and unfamiliarity with healthcare systems in host countries, which led to an inability of expressing their health concerns to healthcare professionals and not knowing health care access procedures, were indicated as influencing their intention to access HIV care services. These seem to reflect a low level of educational attainment as a bigger underlying factor that contributed to these barriers and also indicate a lack of or poor pre-departure preparation, training and information provided by designated agencies or government institutions in the study settings for migrant workers. These findings are consistent with previous findings which found that language difficulties and a lack of knowledge and education about HIV and the availability of HIV care services and unfamiliarity with healthcare systems in host countries were barriers that impede the access of MLHIV to ART or HIV care services in general (18, 23, 26-29, 47). Thus, it is plausible to argue that language barriers result in a lack of self-confidence of migrant workers living with HIV to access health care services they needed, and caused poor health outcomes due to untreated HIV while working abroad. Their untreated HIV may result in reducing the body’s immune system which can contribute to higher risk of acquiring opportunistic infections or co-infections. The status of being undocumented migrant workers, which led to a fear of getting caught by authorities and of being jailed or deported, was also another important barrier that hindered some participants’ access to HIV care services in host countries, which is in line with findings of previous studies reported elsewhere (9, 21-24). The fact that there are undocumented Indonesians working abroad seems to also reflect both poor distribution and access to necessary information about procedures and requirements to become a migrant worker and poor recruitment process of potential migrant workers in the study settings. These issues could be the underlying reasons that influenced people’s decision to be undocumented migrant workers, which ultimately impeded their access to healthcare services while working abroad.

Our study also reported several novel findings which have not been explored in previous studies involving MLHIV (8, 9). We found that the unavailability of HIV care services in the areas where they worked (e.g., plantation areas) and a lack of public transportation between workplaces and healthcare facilities created a significant barrier for some participants to access the services. One of the main reasons was that such conditions may require them to spend a night or more in another place or city where the services are available, resulting in personal outlays of time and money. These considerations result in weighing options between personal finance/time off from work, and accessing HIV services, especially when lacking a social or community support system such as friends who could provide lodging or other support. Similarly, it can be argued that some migrant workers anticipated difficulties acquiring permission from their supervisors or employers to be absent from work, combined with the risk of disclosing their HIV status and the negative sequelae of consequences leading to potential job termination, reporting, workplace stigma, or others. Although stigma and discrimination were not specifically identified as a primary concern of the participants in this study, previous investigations have reported that stigma and discrimination were experienced by MLHIV as barriers that hampered their access to HIV care services (18, 20, 31, 48, 49). Our findings also identified that the mobile, transient nature of their work across districts made it difficult for them to access HIV care services, which has not been reported in previous studies with MLHIV (8, 9). It is apparent that to fully understand HIV care access procedures and systems in different areas requires intensive effort, which seemed challenging for participants in this study who faced language barriers and have limited knowledge and low levels of education.

The use of packaged herbal medicines and traditional medicines, which was a barrier to the migrant workers’ access to HIV care services, was another novel finding of the current study. This seems to be underpinned by some supporting dimensions of accessibility to these medicines, such as the availability and affordability of these medicines (50, 51). These accessibility dimensions were reflected in the fact that participants could afford to buy herbal medicines at public markets or via online sources, or were able to make traditional medicines themselves out of roots, leaves and bark of plants which they could easily find, or via care sought from local traditional healers. Although there have never been studies exploring the influence of packaged herbal medicines and traditional medicines on the access of MLHIV to HIV care services or treatment, some previous studies involving people living with HIV (PLHIV) in general have suggested that the use of traditional medicines has influenced adherence to ART (52-55). Our findings also suggest that peer influence played an important role in the use of these packaged herbal and traditional medicines by the participants. Such influences are reflected in their encouragement for participants to use these medicines for HIV treatment as they are considered more effective than antiretroviral medicines and in the provision of misleading information about the possible negative impacts (e.g., getting caught by authorities, being jailed or deported) of the disclosure of the participants’ HIV status to healthcare professionals. These seem to lead to the participants hiding their HIV status and prevent their access to HIV care services. This current study contributes to the evidence base from previous studies with PLHIV which suggests that social influence among peers is a barrier to accessing proper HIV care services from official health care facilities (56-58).

### 4.1 Study limitations and strengths

We recognise that the use of the snowball sampling technique to recruit participants and the recruitment point at healthcare facilities were limitations of the study as these led to the recruitment of participants from the same networks of the current participants who were already on ART. Thus, we did not include PLHIV who were disengaged from care, as they may have had different stories to share about barriers to their access to HIV care services during their work overseas. This is the first qualitative inquiry exploring barriers to access to HIV care services among Indonesian migrant workers living with HIV. Therefore, our findings are useful to inform government policy makers to address the issues facing HIV-positive migrant workers and to support their health needs through the development of evidence-based policies and interventions.

## 5. Conclusions

This study presents several barriers to access to HIV care services among Indonesian male ex-migrant workers in host countries. The barriers included a lack of language proficiency, a lack of knowledge about healthcare systems in host countries, being undocumented migrant workers, the unavailability of the services in workplaces, long-distance travel to healthcare facilities and a lack of public transportation. The mobile nature of the work which required them to regularly move from one place to another, the use of packaged herbal medicines and traditional medicines, and social influence were also factors that influenced the participants’ access to the services. The findings indicate the need for the dissemination of information and training for migrant workers prior to departure abroad regarding healthcare systems and access procedures in host countries to ensure that they are equipped with necessary information and are aware of how to access healthcare services. The findings also indicate the need for both law and regulation enforcement to prevent undocumented people from going abroad to be migrant workers and to ensure that people are eligible by law to become migrant workers and have the rights to access social and health support in host countries. Future large-scale studies to understand barriers to access to HIV care services by migrant workers living with HIV are recommended.

## Data Availability

All relevant data are within the manuscript.

## Acknowledgements

We would like to thank the participants who had spent their time to voluntarily take part in the interview and provided us with valuable information.

## Supporting information

S1 Fig.: COREQ checklist

