## Supplemental file for "Barriers to access to HIV care services in host countries: views and experiences of Indonesian male ex-migrant workers living with HIV"

Table 1 Consolidated criteria for reporting qualitative studies (COREQ): 32-item checklist

| **No** | **Item** | **Guide questions/description** | Page |
| --- | --- | --- | --- |
| **Domain 1: Research team and reflexivity** | | | |
| Personal Characteristics | | | |
| 1. | Interviewer/facilitator | Which author/s conducted the interview or focus group? | 5 |
| 2 | Credentials | What were the researcher's credentials? E.g. PhD, MD | 5 |
| 3 | Occupation | What was their occupation at the time of the study? | 5 |
| 4 | Gender | Was the researcher male or female? | 5 |
| 5 | Experience and training | What experience or training did the researcher have? | 5 |
| Relationship with participants | | | |
| 6 | Relationship established | Was a relationship established prior to study commencement? | 5 |
| 7 | Participant knowledge of the interviewer | What did the participants know about the researcher? e.g. personal goals, reasons for doing the research | 6 |
| 8 | Interviewer characteristics | What characteristics were reported about the interviewer/facilitator? e.g. Bias, assumptions, reasons and interests in the research topic | 6 |
| **Domain 2: study design** | | | |
| Theoretical framework | | | |
| 9 | Methodological orientation and Theory | What methodological orientation was stated to underpin the study? e.g. grounded theory, discourse analysis, ethnography, phenomenology, content analysis | 5 |
| Participant selection | | | |
| 10 | Sampling | How were participants selected? e.g. purposive, convenience, consecutive, snowball | 5 |
| 11 | Method of approach | How were participants approached? e.g. face-to-face, telephone, mail, email | 5 |
| 12 | Sample size | How many participants were in the study? | 5 |
| 13 | Non-participation | How many people refused to participate or dropped out? Reasons? | 5 |
| Setting | | | |
| 14 | Setting of data collection | Where was the data collected? e.g. home, clinic, workplace | 6 |
| 15 | Presence of non-participants | Was anyone else present besides the participants and researchers? | 6 |
| 16 | Description of sample | What are the important characteristics of the sample? e.g. demographic data, date | 6 |
| Data collection | | | |
| 17 | Interview guide | Were questions, prompts, guides provided by the authors? Was it pilot tested? | 6 |
| 18 | Repeat interviews | Were repeat interviews carried out? If yes, how many? | 7 |
| 19 | Audio/visual recording | Did the research use audio or visual recording to collect the data? | 6 |
| 20 | Field notes | Were field notes made during and/or after the interview or focus group? | 6 |
| 21 | Duration | What was the duration of the interviews or focus group? | 6 |
| 22 | Data saturation | Was data saturation discussed? | 6-7 |
| 23 | Transcripts returned | Were transcripts returned to participants for comment and/or correction? | 7 |
| **Domain 3: analysis and findings** | | | |
| Data analysis | | | |
| 24 | Number of data coders | How many data coders coded the data? | 7 |
| 25 | Description of the coding tree | Did authors provide a description of the coding tree? | - |
| 26 | Derivation of themes | Were themes identified in advance or derived from the data? | 7 |
| 27 | Software | What software, if applicable, was used to manage the data? | 7 |
| 28 | Participant checking | Did participants provide feedback on the findings? | 7 |
| Reporting | | | |
| 29 | Quotations presented | Were participant quotations presented to illustrate the themes / findings? Was each quotation identified? e.g. participant number | 8-13 |
| 30 | Data and findings consistent | Was there consistency between the data presented and the findings? | 8-13 |
| 31 | Clarity of major themes | Were major themes clearly presented in the findings? | 8-13 |
| 32 | Clarity of minor themes | Is there a description of diverse cases or discussion of minor themes? | 8-13 |
